# Differential symptoms among COVID-19 outpatients before and during periods of SARS-CoV-2 Omicron variant dominance in Blantyre, Malawi: a prospective observational study

**DOI:** 10.1101/2022.07.15.22277665

**Authors:** Marah G. Chibwana, Herbert W. Thole, Cat Anscombe, Philip M. Ashton, Edward Green, Kayla G. Barnes, Jen Cornick, Ann Turner, Desiree Witte, Sharon Nthala, Chikondi Thom, Felistas Kanyandula, Anna Ainani, Natasha Mtike, Hope Tambala, Veronica N’goma, Dorah Mwafulirwa, Erick Asima, Ben Morton, Markus Gmeiner, Zaziwe Gundah, Gift Kawalazira, Neil French, Nicholas Feasey, Robert S. Heyderman, Todd D Swarthout, Kondwani C. Jambo

## Abstract

**Background:** It is widely reported that the SARS-CoV-2 Omicron variant has resulted in high number of cases, but relatively low incidence of severe disease and deaths, compared to the pre-Omicron variants of concern. We aim to assess the differences in symptom prevalence between Omicron and pre-Omicron infections in a sub-Saharan African population.

**Methods:** In this cross-sectional observational study, we collected data from children and adult outpatients presenting at two primary healthcare facilities in Blantyre, Malawi. Eligible participants were aged >1month old, with signs suggestive of COVID-19, and those not suspected of COVID-19. Nasopharyngeal swabs were collected for SARS-CoV-2 PCR testing and positive samples whole genome sequenced to identify the infecting variant. The primary outcome was the likelihood of presenting with a given symptom in individuals testing positive during the period in which Omicron-dominated (December 2021 to March 2022) with those infected during the pre-Omicron period (August 2021 to November 2021).

**Findings:** Among 5176 study participants, the median age was 28 years (IQR 21-38), of which 6.4% were under 5, 9.2% were 6 to 17 years, 77% were 18 to 50 years, and 7.1% were above 50 years old. Prevalence of SARS-CoV-2 infection was 23% (1187/5188), varying over time, with peaks in January 2021, July 2021 and December 2021, driven by the Beta (B.1.351), Delta (B.1.617.2) and Omicron (BA.1/2) variants, respectively. Headache (OR 0.47[CI 0.29 – 0.79]), cough (OR 0.37[CI 0.22 – 0.61]), fatigue (OR 0.20[CI 0.08 – 0.48]) and abdominal pain (OR 0.38[CI 0.18 – 0.78]) were less common in participants infected during the Omicron-dominant period than during pre-Omicron period. Fever was more common in participants infected during the Omicron-dominated period than during pre-Omicron period (OR 2.46[CI 1.29 – 4.97]). COVID-19 vaccination, accounting for number of doses and days since last dose, was not associated with a reduced risk of PCR-confirmed SARS-CoV-2 infection (1 dose, OR 1.10[CI 0.39 – 2.66]; 2 doses, OR 1.11[CI 0.40 – 2.57]; all p=0.8).

**Interpretation:** In this Malawian population, the prevalence of clinical symptoms associated with Omicron infection differ from those of pre-Omicron infections and may be harder to identify clinically with current symptom guidelines. To maintain robust surveillance for COVID-19 and emerging variants, case definitions and testing policies will need to be regularly reviewed to ensure case ascertainment.

## INTRODUCTION

As of June 2022, the COVID-19 pandemic has resulted in 543 million cases and 6.3 million deaths globally ^1^. In sub-Saharan Africa, the pandemic has however been associated with a lower rate of hospitalisation and deaths than in Europe and the Americas ^1^, despite widespread SARS-CoV-2 community transmission ^2^, and low COVID-19 vaccine coverage ^3^. Due to the continued emergence of SARS-CoV-2 variants of concern, surveillance is essential for monitoring the pandemic and informing public health interventions, however the optimal approach to surveillance in low-income, resource-poor settings is unclear ^4^.

By June 2022, Malawi had experienced four epidemic waves peaking in July 2020, January 2021, July 2021, and December 2021. There were 86,348 confirmed SARS-CoV-2 cases nationally, with 2,645 COVID-19-associated deaths ^1^. However, due to the availability of testing there is considerable case under ascertainment, as evidence by the high seroprevalence of >65% observed in Malawi as of July 2021 ^5^. Blantyre has had the highest number of reported COVID-19 cases in Malawi, with 28.6% of the national cases ^6^. Recently, the Omicron variant has resulted in less hospitalisations and mortality in Malawi compared to the Delta variant ^1^, which has coincided with high seroprevalence of SARS-CoV-2 antibodies in Malawi and across sub-Saharan Africa ^5,7^. Further, Malawi introduced COVID-19 vaccines in March 2021 ^8^. The COVID-19 vaccination coverage for Malawi is 4.5%, including the AstraZeneca, Janssen and Pfizer vaccines, with AstraZeneca vaccine constituting most of the doses ^8^.

Using data from early in the pandemic, a standardised case definition for COVID-19 was developed by the World Health Organisation (WHO) ^9^ and United States Centre for Disease Control (US CDC) ^10^, and these have allowed targeted SARS-CoV-2 testing. However, there has been limited systematic data collected on clinical presentation of SARS-CoV-2 infection in sub-Saharan African settings, outside of South Africa. This is particularly important as there is a high burden of other febrile illnesses such as malaria, pneumonia, TB and salmonellosis that have clinical features that overlap with COVID-19 ^11,12^. Further, there is limited data on the differences in clinical presentation between infections caused by different variants of concern (VOC), especially amongst non-hospitalised patients. To address these gaps, our study measured the prevalence of PCR-confirmed SARS-CoV-2 infection among outpatients presenting with medical conditions at primary healthcare facilities and compared the symptom profiles between Omicron and pre-Omicron infections.

## METHODS

### Study design and population

From November 2020 to March 2022, we conducted a SARS-CoV-2 prevalence study in primary healthcare facilities in the city of Blantyre, southern Malawi. Blantyre is Malawi’s commercial city with a population of 800,264 (pop density, 3334/km^2^). Adults and children were recruited voluntarily from two government-owned primary healthcare facilities, Ndirande Health Centre and Limbe Health Centre, both overseen by the Blantyre District Health Office. Census data shows Ndirande HC serves a catchment area of 135,736, while Limbe HC serves a catchment area of 145,604, but the actual catchment is likely much higher.

From November 2020 to July 2021, individuals with medical conditions were screened at the facility’s outpatient services department. Following assessment by a clinician and a review of the individual’s health passport (patient retained medical record), a nasopharyngeal swab was collected for SARS-CoV-2 screening by SARS-CoV-2 by RT-PCR from patients suspected of COVID-19 according to the WHO case definition ^9^. From August 2021, following a protocol amendment to include capturing a more detailed clinical history using the ISARIC symptom list ^13-15^, participants included both those suspected and those not suspected of COVID-19 according to the WHO case definition, with a 2:1 numerical bias towards those with suspected COVID-19. Patients with suspected COVID-19 were recruited as and when they presented to the facility, while those with not clinically suspected of COVID-19 were selected by approaching every third patient in health facility’s triage area.

### Data and specimen collection

Following informed consent and assent (for children) from August 2021, we used an abridged International Severe Acute Respiratory and Emerging Infection Consortium (ISARIC) Clinical Characterisation Protocol (CCP) electronic case report form (eCRF) ^13-15^ to collect demographic and clinical data from all participants. Study nurses collected nasopharyngeal swabs in Universal Transport Medium (UTM) (Copan, Brescia, Italy) from all participants. Samples were initially stored and transported to the Malawi-Liverpool-Wellcome Programme (MLW) laboratory on ice and processed within 48 hours.

### Laboratory testing

Nasopharyngeal swabs were tested for SARS-CoV-2 RNA using the CDC 2019-nCoV RNA RT-PCR diagnostic panel (Integrated DNA Technologies, Iowa, USA). A cycle threshold (Ct) value of <40 was considered positive for SARS-CoV-2 using QuantStudio Real-Time PCR software v1.3 (Applied Biosystems, UK). Ribonuclease protein was used as an internal control to identify presence of human RNA. A negative extraction control and a PCR no-template control were also performed with every test. The results of patients with positive PCR tests were shared with the Blantyre District Health Office for further follow up and patient management.

### Genomic sequencing and analysis

Samples were extracted using the Qiasymphony-DSP mini kit 200 (Qiagen, UK) with offboard lysis. Samples were then tested using the CDC N1 assay to confirm the Ct values before sequencing. Samples with a Ct value <27 were sequenced. The following sequencing protocols were used; ARTICv2 and v3 was used from November 2020 from July 2021 to July 2021 ^16^ and UNZA ^17^ from August 2021 on wards. Initially two primer pools were used, however a third pool was made for primer pairs that commonly had lower depth compared to the average ^15^. PCR cycling conditions were adapted to the new sequencing primers, with annealing temperature changed to 60°C. Sequencing was carried out with the Oxford Nanopore Technologies MinION sequencer. Samples that had poor coverage (<70%) with the ARTIC primer set were repeated with the UNZA primer set.

For analysis of sequencing data, the lineage of each consensus genome was identified using pangolin with the following versions; pangolin v3.1.17, pangolearn 2021-12-06, constellations v0.1.1, scorpio v0.3.16, pango-designation used by pangoLEARN/Usher v1.2.105, pango-designation aliases v1.2.122 ^18^. Samples were re-analysed when the Pangolin database was updated. The run was repeated if there was contamination in the negative control.

### Statistical analysis

We performed statistical analyses and graphical presentation using R statistical package, version 4.1.0. Categorical variables were summarised using frequency distributions and compared using Pearson’s Chi-squared test and Fisher’s exact test. The continuous variables were presented as median with interquartile range.

We employed multivariable logistic regression models, as implemented in the R package stats (version 3.6.2), to investigate odds of presenting with particular symptoms in Omicron compared pre-Omicron phases, adjusting for age and sex. A multivariable logistic regression model adjusting for age, sex, vaccination status, vaccination doses and days since last vaccine dose was also employed to investigate the impact of COVID-19 vaccination on PCR-confirmed SARS-CoV-2. P-values <0.05 were considered significant.

### Ethics approval

The study was approved by the College of Medicine Research and Ethics Committee (COMREC P.08/20/3099) and Liverpool School of Tropical Medicine Research Ethics Committee (LSTMREC 21-058).

## RESULTS

### Demographic and clinical characteristics of participants

From November 2020 through March 2022, 6147 (Ndirande Health Centre, n=2899; Limbe Health Centre, n=3248) individuals were approached, and 5188 participants were enrolled but 12 were excluded as they did not have sex recorded. Refusals were mostly from parents or guardians who did not consent for their children to undergo nasopharyngeal swabbing, and this did not change throughout the study period. Overall, the participants’ median age was 28 years (IQR 21-38), of which 6.4% (331/5176) were under 5 years, 9.2% (331/5176) were 6 to 17 years (479/5176), 77% (4000/5176) were 18 to 50 years, and 7.1% (368/5176) were above 50 years old. Of the total 50% (2596/5176) were female (**table 1**).

**Table 1.** Participant characteristics for main cohort.

| Characteristic | N = 5,176<sup>†</sup> |
| --- | --- |
| **Sex** |  |
| Female | 2,596 (50%) |
| Male | 2,580 (50%) |
| **Age** | 28 (21, 38) |
| **Age group** |  |
| 1. 0-5 years | 331 (6.4%) |
| 2. 6-12 years | 228 (4.4%) |
| 3. 13-17 years | 249 (4.8%) |
| 4. 18-50 years | 4,000 (77%) |
| 5. 51+ years | 368 (7.1%) |
| **SARS-CoV-2 PCR positivity** |  |
| Negative | 3,992 (77%) |
| Positive | 1,184 (23%) |
| <sup>†</sup> n (%); Median (IQR) |  |

### Prevalence of SARS-CoV-2 infection and genomic surveillance

The overall prevalence of PCR-confirmed SARS-CoV-2 infection was 23% (1187/5176) (**table 1**). SARS-CoV-2 prevalence varied over time, with three distinct peaks over the study period, namely January 2021, July 2021, and December 2021 (**figure 1A**). SARS-CoV-2 prevalence was lowest in those under 5 years of age (5.74% [CI 3.49 – 8.82]) compared to all other age groups (6-12yrs, 16.7% [12.1 – 22.2]; 13-17yrs, 25.3% [20.0 – 31.2]; 18-50yrs, 24.5% [23.2 – 25.9]; 50+yrs, 22.8% [18.6 −27.5) (**figure 1B**). The three prevalence peaks corresponded with emergence of variants of concern (VOC), including Beta (B.1.351; January 2021), Delta (1.617.2; July 2021) and Omicron (BA.1/2; December 2021) (**figure 1C**). Only Omicron (BA.1) was detectable in all age groups (**figure 1D**).

**Figure 1.**
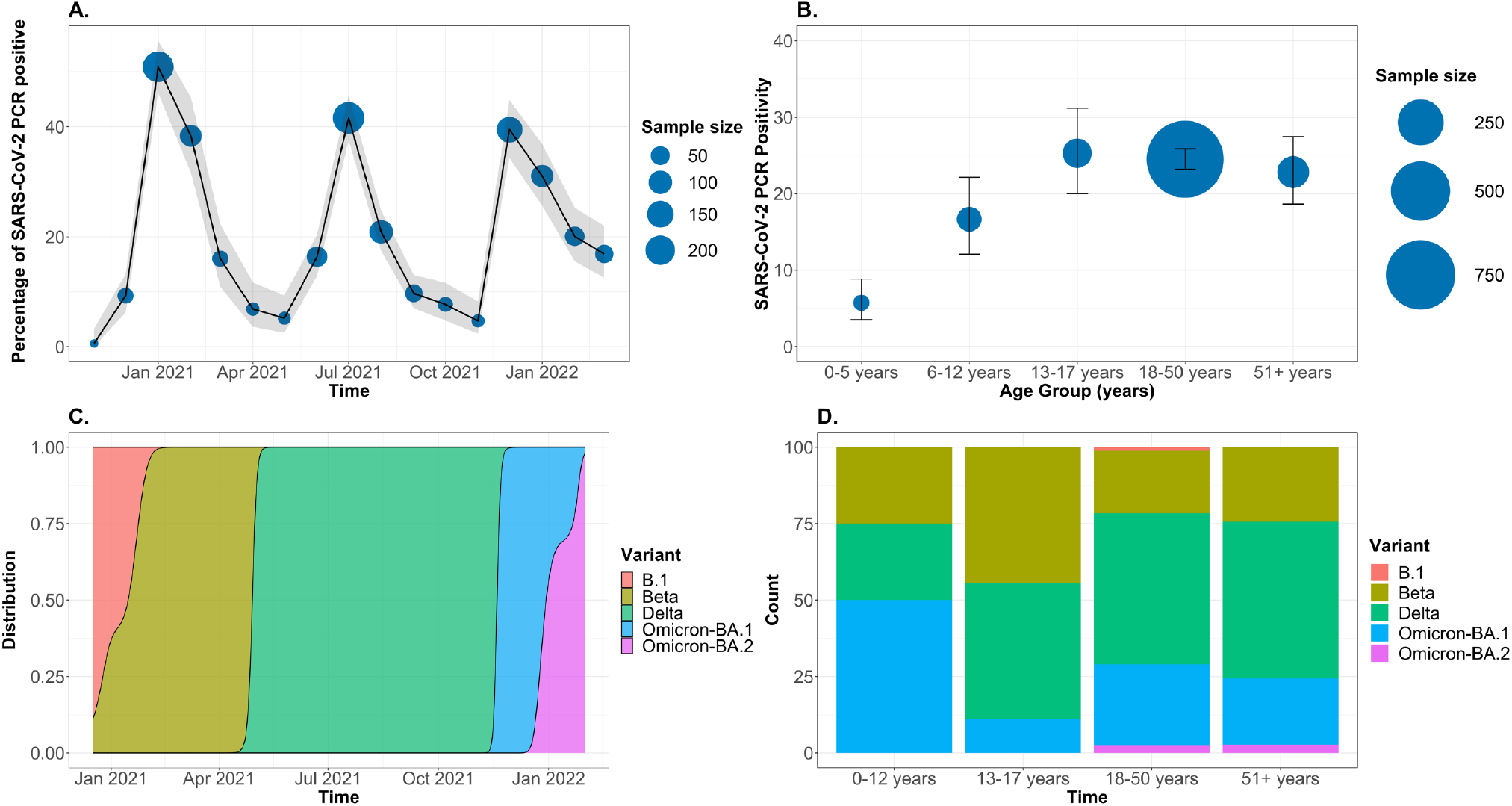
Prevalence of SARS-CoV-2 infections. A) Prevalence of SARS-CoV-2 infections across age groups. B) Prevalence of SARS-CoV-2 infections over time. Grey area represents confidence intervals. C) SARS-CoV-2 variants of concern across the three pandemic waves. D) SARS-CoV-2 variants of concern across age groups. (n=402)

### Symptoms associated with SARS-CoV-2 infection, pre- and during the Omicron-dominated phase

Forty-nine percent (2520/5176) of the total participants were recruited from August 2021 to March 2022, hence had detailed symptom and medical history (**Supplementary figure 1**). Out of these 2509 had complete symptomology data and were used in the subsequent analysis. Using a multivariable analysis, cough (71% vs 68%, risk ratio, 1.50; 95% CI, 1.00 to 2.30, p=0.056), fatigue (14% vs 6.3%, risk ratio, 2.27; 95% CI, 1.29 to 3.86, p=0.003) and headache (49% vs 37%, risk ratio, 1.64; 95% CI, 1.15 to 2.34, p=0.007) were associated with a high risk of PCR-confirmed SARS-CoV-2 infection during the pre-Omicron period (**table 2**). While, during the Omicron-dominated period, only headache (39% vs 30%, risk ratio, 1.41; 95% CI, 1.07 to 1.86, p=0.015) was associated with a high risk of PCR-confirmed SARS-CoV-2 infection.

**Table 2.** Symptoms associated with SARS-CoV-2 infection.

| Characteristic | SARS-CoV-2 PCR Result (Pre-Omicron) |  |  |  | Univariate (Pre-Omicron) |  |  |  | Multivariable (Pre-Omicron) |  |  | SARS-CoV-2 PCR Result (Omicron) |  |  |  | Univariate (Omicron) |  |  |  | Multivariable (Omicron) |  |
| --- | --- | --- | --- | --- | --- | --- | --- | --- | --- | --- | --- | --- | --- | --- | --- | --- | --- | --- | --- | --- | --- |
|  | Overall, N = 1,311<sup>1</sup> | Negative, N = 1,157<sup>1</sup> | Positive, N = 154<sup>1</sup> | p-value<sup>2</sup> | N | OR<sup>3</sup> | 95% CI<sup>3</sup> | p-value | OR<sup>3</sup> | 95% CI<sup>3</sup> | p-value | Overall, N = 1,198<sup>1</sup> | Negative, N = 870<sup>1</sup> | Positive, N = 328<sup>1</sup> | p-value<sup>2</sup> | N | OR<sup>3</sup> | 95% CI<sup>3</sup> | p-value | OR<sup>3</sup> | 95% CI<sup>3</sup> |
| Fever | 229 (17%) | 207 (18%) | 22 (14%) | 0.3 | 1,311 |  |  |  |  |  |  | 217 (18%) | 148 (17%) | 69 (21%) | 0.11 | 1,198 |  |  |  |  |  |
| No |  |  |  |  |  | Ref. | — |  | Ref. | — |  |  |  |  |  |  | Ref. | — |  | Ref. | — |
| Yes |  |  |  |  |  | 0.76 | 0.46, 1.21 | 0.3 | 0.65 | 0.39, 1.05 | 0.093 |  |  |  |  |  | 1.30 | 0.94, 1.78 | 0.11 | 1.23 | 0.88, 1.71 |
| Cough | 893 (68%) | 784 (68%) | 109 (71%) | 0.5 | 1,311 |  |  |  |  |  |  | 654 (55%) | 477 (55%) | 177 (54%) | 0.8 | 1,198 |  |  |  |  |  |
| No |  |  |  |  |  | Ref. | — |  | Ref. | — |  |  |  |  |  |  | Ref. | — |  | Ref. | — |
| Yes |  |  |  |  |  | 1.15 | 0.80, 1.68 | 0.5 | 1.50 | 1.00, 2.30 | **0.056** |  |  |  |  |  | 0.97 | 0.75, 1.25 | 0.8 | 1.04 | 0.79, 1.38 |
| Fatigue | 94 (7.2%) | 73 (6.3%) | 21 (14%) | **<0.001** | 1,311 |  |  |  |  |  |  | 32 (2.7%) | 21 (2.4%) | 11 (3.4%) | 0.4 | 1,198 |  |  |  |  |  |
| No |  |  |  |  |  | Ref. | — |  | Ref. | — |  |  |  |  |  |  | Ref. | — |  | Ref. | — |
| Yes |  |  |  |  |  | 2.34 | 1.37, 3.87 | **0.001** | 2.27 | 1.29, 3.86 | **0.003** |  |  |  |  |  | 1.40 | 0.65, 2.89 | 0.4 | 1.24 | 0.55, 2.64 |
| Loss of smell | 62 (4.7%) | 53 (4.6%) | 9 (5.8%) | 0.5 | 1,311 |  |  |  |  |  |  | 25 (2.1%) | 16 (1.8%) | 9 (2.7%) | 0.3 | 1,198 |  |  |  |  |  |
| No |  |  |  |  |  | Ref. | — |  | Ref. | — |  |  |  |  |  |  | Ref. | — |  | Ref. | — |
| Yes |  |  |  |  |  | 1.29 | 0.59, 2.55 | 0.5 | 0.89 | 0.34, 2.16 | 0.8 |  |  |  |  |  | 1.51 | 0.63, 3.38 | 0.3 | 0.70 | 0.22, 2.06 |
| Loss of taste | 86 (6.6%) | 70 (6.1%) | 16 (10%) | **0.041** | 1,311 |  |  |  |  |  |  | 37 (3.1%) | 21 (2.4%) | 16 (4.9%) | **0.028** | 1,198 |  |  |  |  |  |
| No |  |  |  |  |  | Ref. | — |  | Ref. | — |  |  |  |  |  |  | Ref. | — |  | Ref. | — |
| Yes |  |  |  |  |  | 1.80 | 0.99, 3.11 | **0.044** | 1.79 | 0.83, 3.65 | 0.12 |  |  |  |  |  | 2.07 | 1.05, 4.01 | **0.031** | 2.04 | 0.81, 5.13 |
| Sore throat | 154 (12%) | 134 (12%) | 20 (13%) | 0.6 | 1,311 |  |  |  |  |  |  | 161 (13%) | 108 (12%) | 53 (16%) | 0.090 | 1,198 |  |  |  |  |  |
| No |  |  |  |  |  | Ref. | — |  | Ref. | — |  |  |  |  |  |  | Ref. | — |  | Ref. | — |
| Yes |  |  |  |  |  | 1.14 | 0.67, 1.85 | 0.6 | 1.03 | 0.59, 1.72 | >0.9 |  |  |  |  |  | 1.36 | 0.95, 1.93 | 0.091 | 1.35 | 0.93, 1.93 |
| Headache | 501 (38%) | 426 (37%) | 75 (49%) | **0.004** | 1,311 |  |  |  |  |  |  | 389 (32%) | 261 (30%) | 128 (39%) | **0.003** | 1,198 |  |  |  |  |  |
| No |  |  |  |  |  | Ref. | — |  | Ref. | — |  |  |  |  |  |  | Ref. | — |  | Ref. | — |
| Yes |  |  |  |  |  | 1.63 | 1.16, 2.28 | **0.005** | 1.64 | 1.15, 2.34 | **0.007** |  |  |  |  |  | 1.49 | 1.14, 1.95 | **0.003** | 1.41 | 1.07, 1.86 |
| Muscle ache | 270 (21%) | 233 (20%) | 37 (24%) | 0.3 | 1,311 |  |  |  |  |  |  | 343 (29%) | 235 (27%) | 108 (33%) | **0.043** | 1,198 |  |  |  |  |  |
| No |  |  |  |  |  | Ref. | — |  | Ref. | — |  |  |  |  |  |  | Ref. | — |  | Ref. | — |
| Yes |  |  |  |  |  | 1.25 | 0.83, 1.85 | 0.3 | 1.15 | 0.72, 1.80 | 0.6 |  |  |  |  |  | 1.33 | 1.01, 1.74 | **0.044** | 1.28 | 0.94, 1.73 |
| Joint pain | 105 (8.0%) | 89 (7.7%) | 16 (10%) | 0.2 | 1,311 |  |  |  |  |  |  | 75 (6.3%) | 49 (5.6%) | 26 (7.9%) | 0.14 | 1,198 |  |  |  |  |  |
| No |  |  |  |  |  | Ref. | — |  | Ref. | — |  |  |  |  |  |  | Ref. | — |  | Ref. | — |
| Yes |  |  |  |  |  | 1.39 | 0.77, 2.38 | 0.2 | 1.07 | 0.54, 2.01 | 0.8 |  |  |  |  |  | 1.44 | 0.87, 2.34 | 0.15 | 1.00 | 0.57, 1.73 |
| Abdominal pain | 159 (12%) | 137 (12%) | 22 (14%) | 0.4 | 1,311 |  |  |  |  |  |  | 96 (8.0%) | 68 (7.8%) | 28 (8.5%) | 0.7 | 1,198 |  |  |  |  |  |
| No |  |  |  |  |  | Ref. | — |  | Ref. | — |  |  |  |  |  |  | Ref. | — |  | Ref. | — |
| Yes |  |  |  |  |  | 1.24 | 0.75, 1.98 | 0.4 | 1.33 | 0.76, 2.23 | 0.3 |  |  |  |  |  | 1.10 | 0.69, 1.72 | 0.7 | 1.29 | 0.79, 2.08 |
| Diarrhoea | 102 (7.8%) | 92 (8.0%) | 10 (6.5%) | 0.5 | 1,311 |  |  |  |  |  |  | 82 (6.8%) | 63 (7.2%) | 19 (5.8%) | 0.4 | 1,198 |  |  |  |  |  |
| No |  |  |  |  |  | Ref. | — |  | Ref. | — |  |  |  |  |  |  | Ref. | — |  | Ref. | — |
| Yes |  |  |  |  |  | 0.80 | 0.39, 1.51 | 0.5 | 0.89 | 0.41, 1.78 | 0.8 |  |  |  |  |  | 0.79 | 0.45, 1.31 | 0.4 | 0.87 | 0.49, 1.49 |
| Shortness of breath | 108 (8.2%) | 100 (8.6%) | 8 (5.2%) | 0.14 | 1,311 |  |  |  |  |  |  | 86 (7.2%) | 66 (7.6%) | 20 (6.1%) | 0.4 | 1,198 |  |  |  |  |  |
| No |  |  |  |  |  | Ref. | — |  | Ref. | — |  |  |  |  |  |  | Ref. | — |  | Ref. | — |
| Yes |  |  |  |  |  | 0.58 | 0.25, 1.14 | 0.15 | 0.55 | 0.23, 1.15 | 0.14 |  |  |  |  |  | 0.79 | 0.46, 1.30 | 0.4 | 0.80 | 0.41, 1.52 |
| Chest pain | 244 (19%) | 217 (19%) | 27 (18%) | 0.7 | 1,311 |  |  |  |  |  |  | 156 (13%) | 108 (12%) | 48 (15%) | 0.3 | 1,198 |  |  |  |  |  |
| No |  |  |  |  |  | Ref. | — |  | Ref. | — |  |  |  |  |  |  | Ref. | — |  | Ref. | — |
| Yes |  |  |  |  |  | 0.92 | 0.58, 1.41 | 0.7 | 0.83 | 0.51, 1.32 | 0.5 |  |  |  |  |  | 1.21 | 0.83, 1.74 | 0.3 | 1.32 | 0.90, 1.93 |
| Runny nose | 416 (32%) | 372 (32%) | 44 (29%) | 0.4 | 1,311 |  |  |  |  |  |  | 261 (22%) | 191 (22%) | 70 (21%) | 0.8 | 1,198 |  |  |  |  |  |
| No |  |  |  |  |  | Ref. | — |  | Ref. | — |  |  |  |  |  |  | Ref. | — |  | Ref. | — |
| Yes |  |  |  |  |  | 0.84 | 0.58, 1.21 | 0.4 | 0.78 | 0.52, 1.16 | 0.2 |  |  |  |  |  | 0.96 | 0.70, 1.31 | 0.8 | 0.96 | 0.69, 1.32 |
| Pneumonia | 80 (6.1%) | 70 (6.1%) | 10 (6.5%) | 0.8 | 1,311 |  |  |  |  |  |  | 72 (6.0%) | 57 (6.6%) | 15 (4.6%) | 0.2 | 1,198 |  |  |  |  |  |
| No |  |  |  |  |  | Ref. | — |  | Ref. | — |  |  |  |  |  |  | Ref. | — |  | Ref. | — |
| Yes |  |  |  |  |  | 1.08 | 0.51, 2.05 | 0.8 | 1.51 | 0.67, 3.13 | 0.3 |  |  |  |  |  | 0.68 | 0.37, 1.19 | 0.2 | 0.78 | 0.37, 1.58 |
| <sup>1</sup> n (%) |  |  |  |  |  |  |  |  |  |  |  |  |  |  |  |  |  |  |  |  |  |
| <sup>2</sup> Pearson's Chi-squared test |  |  |  |  |  |  |  |  |  |  |  |  |  |  |  |  |  |  |  |  |  |
| <sup>3</sup> OR = Odds Ratio. CI = Confidence Interval |  |  |  |  |  |  |  |  |  |  |  |  |  |  |  |  |  |  |  |  |  |

<sup>1</sup> n (%)
<sup>2</sup> Pearson's Chi-squared test
<sup>3</sup> OR = Odds Ratio, CI = Confidence Interval

### Impact of COVID-19 vaccination on the risk of PCR-confirmed SARS-CoV-2 infection

Eighty percent (2009/2520) of the participants with detailed symptomology were eligible for vaccination (18 years and above) and had complete vaccination history. We, therefore, used these individuals to determine whether the risk of PCR-confirmed SARS-CoV-2 infection was different between vaccinated compared to unvaccinated adults. Using a multivariable analysis, adjusting for days since last vaccine dose, sex, age and recruitment period; COVID-19 vaccination was not associated with a reduced risk of PCR-confirmed SARS-CoV-2 infection (1 dose, OR 1.10[CI 0.39-2.66]; 2 doses, OR 1.11[CI 0.40-2.57]; <91 days, OR 1.93[CI 0.74-5.68]; 91+ days, OR 1.54[CI 0.63-4.34]) (**table 3**). However, the recruitment period of December 2021 to March 2022 (Omicron), was associated with a threefold increase in the risk of PCR-confirmed SARS-CoV-2 infection than the August 2021 to November 2021 (pre-Omicron) period (OR 2.56 [CI 2.02-3.26]) (**table 3**).

**Table 3.** Participant characteristics for PCR-confirmed SARS-CoV-2 infected adult patients.

| Characteristic | Overall, N = 447<sup>1</sup> | Omicron phase, N = 302<sup>1</sup> | Pre-omicron phase, N = 145<sup>1</sup> | p-value<sup>2</sup> |
| --- | --- | --- | --- | --- |
| **sex** |  |  |  | 0.2 |
| Female | 232 (52%) | 151 (50%) | 81 (56%) |  |
| Male | 215 (48%) | 151 (50%) | 64 (44%) |  |
| **Age** | 31 (24, 39) | 31 (25, 38) | 32 (23, 39) | 0.8 |
| **HIV status** |  |  |  | 0.4 |
| Negative | 277 (84%) | 173 (83%) | 104 (87%) |  |
| Positive | 51 (16%) | 35 (17%) | 16 (13%) |  |
| Unknown | 119 | 94 | 25 |  |
| **PLHIV on ART** | 49 (11%) | 34 (11%) | 15 (10%) | 0.8 |
| **Hypertension** | 7 (1.6%) | 5 (1.7%) | 2 (1.4%) | >0.9 |
| **Diabetes** | 4 (0.9%) | 0 (0%) | 4 (2.8%) | **0.011** |
| **Asthma** | 15 (3.4%) | 8 (2.6%) | 7 (4.8%) | 0.3 |
| **COVID-19 vaccination (doses)** |  |  |  | **0.025** |
| 0 | 310 (69%) | 206 (68%) | 104 (72%) |  |
| 1 | 63 (14%) | 37 (12%) | 26 (18%) |  |
| 2 | 74 (17%) | 59 (20%) | 15 (10%) |  |
| **Days since last vaccine dose** | 137 (51, 187) | 173 (141, 208) | 78 (40, 137) | **<0.001** |
| Unknown | 375 | 263 | 112 |  |

<sup>1</sup> n (%); Median (IQR)
<sup>2</sup> Pearson's Chi-squared test; Wilcoxon rank sum test; Fisher's exact test

### Symptoms associated with PCR-confirmed SARS-CoV-2 infection change with variants

Based on the genomic surveillance data (**figure 1C**), we assigned all individuals infected within the period from August 2021 to November 2021 as pre-Omicron infections (most commonly Delta infections), and those from December 2021 to March 2022 as Omicron infections. Twenty two percent (447/2009) of vaccine-eligible patients who had provided the date since last vaccine dose had PCR-confirmed SARS-CoV-2 infection (**table 4**), while 35 out of 228 patients were less than 18 years of age and had PCR-confirmed SARS-CoV-2 infection (**supplementary table 1**), as such subsequent analyses focused on the adult population (18 years and above). Fifty percent (151/302) of these patients in Omicron phase were female, while 56% (81/145) were female in the pre-Omicron phase. The median age of patients from the two phases was similar (31 years [IQR 25-38] vs. 32[IQR 23-39], p=0.8) (**table 4**). Among those with known self-reported HIV status, the HIV prevalence was 16%, which was similar between the two phases (17% vs 13%, p=0.4). Of those People Living with HIV (PLHIV), 97% (34/35) in the Omicron phase and 94% (15/16) in the pre-Omicron phase were on antiretroviral therapy. Other chronic illnesses were uncommon, with only 1.6% (7/447), 0.9% (4/447) and 3.4% (15/447) self-reported to have hypertension, diabetes, and asthma, respectively, although hypertension and diabetes, in particular, are widely underdiagnosed in Malawi ^19,20^. Thirty-one percent (137/447) of the participants reported being vaccinated with at least one dose of the COVID-19 vaccine (AstraZeneca vaccine (80% (109/137)) or Janssen vaccine (20% (28/137)). Of which, 54% (74/447) had self-reported to have received at least two doses of the AstraZeneca vaccine, with more patients in the Omicron phase than the pre-Omicron phase (20% (59/302) vs. 10% (15/145), p=0.025). Moreover, the days since last vaccine dose were longer in the Omicron phase than pre-Omicron phase (173[IQR 141-208] vs. 78[IQR 40-137], p<0.001).

**Table 4.** COVID-19 vaccination and the risk of PCR-confirmed SARS-CoV-2 infection.

| Characteristic | Seropositivity |  |  |  | Univariate |  |  |  | Multivariable |  |  |
| --- | --- | --- | --- | --- | --- | --- | --- | --- | --- | --- | --- |
|  | Overall, N = 2,009<sup>1</sup> | 0, N = 1,627<sup>1</sup> | 1, N = 382<sup>1</sup> | p-value<sup>2</sup> | N | OR<sup>3</sup> | 95% CI<sup>3</sup> | p-value | OR<sup>3</sup> | 95% CI<sup>3</sup> | p-value |
| **COVID-19 vaccine doses** |  |  |  | **0.027** | 2,009 |  |  |  |  |  |  |
| 0 | 1,718 (100%) | 1,408 (82%) | 310 (18%) |  |  | Ref. | — |  | Ref. | — |  |
| 1 | 142 (100%) | 107 (75%) | 35 (25%) |  |  | 1.49 | 0.98, 2.20 | **0.053** | 1.10 | 0.39, 2.66 | 0.8 |
| 2 | 149 (100%) | 112 (75%) | 37 (25%) |  |  | 1.50 | 1.00, 2.20 | **0.042** | 1.11 | 0.40, 2.57 | 0.8 |
| **Days since last vaccine dose** |  |  |  | **0.003** | 2,009 |  |  |  |  |  |  |
| 0 | 1,763 (100%) | 1,447 (82%) | 316 (18%) |  |  | Ref. | — |  | Ref. | — |  |
| <91 days | 90 (100%) | 68 (76%) | 22 (24%) |  |  | 1.48 | 0.88, 2.39 | 0.12 | 1.93 | 0.74, 5.68 | 0.2 |
| 91+ days | 156 (100%) | 112 (72%) | 44 (28%) |  |  | 1.80 | 1.23, 2.58 | **0.002** | 1.54 | 0.63, 4.34 | 0.4 |
| **Sex** |  |  |  | 0.4 | 2,009 |  |  |  |  |  |  |
| Female | 1,001 (100%) | 803 (80%) | 198 (20%) |  |  | Ref. | — |  | Ref. | — |  |
| Male | 1,008 (100%) | 824 (82%) | 184 (18%) |  |  | 0.91 | 0.72, 1.13 | 0.4 | 0.84 | 0.67, 1.06 | 0.15 |
| **Age group** |  |  |  | 0.13 | 2,009 |  |  |  |  |  |  |
| 18–50 years | 1,904 (100%) | 1,536 (81%) | 368 (19%) |  |  | Ref. | — |  | Ref. | — |  |
| 51+ years | 105 (100%) | 91 (87%) | 14 (13%) |  |  | 0.64 | 0.35, 1.10 | 0.13 | 0.62 | 0.33, 1.09 | 0.12 |
| **Recruitment period** |  |  |  | **<0.001** | 2,009 |  |  |  |  |  |  |
| Pre-omicron phase | 1,080 (100%) | 943 (87%) | 137 (13%) |  |  | Ref. | — |  | Ref. | — |  |
| Omicron phase | 929 (100%) | 684 (74%) | 245 (26%) |  |  | 2.47 | 1.96, 3.11 | **<0.001** | 2.56 | 2.02, 3.26 | **<0.001** |

<sup>1</sup> n (%)
<sup>2</sup> Pearson's Chi-squared test
<sup>3</sup> OR = Odds Ratio, CI = Confidence Interval

Clinical symptoms associated with PCR-confirmed SARS-CoV-2 infections were different during the Omicron and pre-Omicron phases (**table 5**). Cough (70% vs. 54%, p<0.001), fatigue (14% vs 3.6%, p<0.001), loss of taste (10% vs. 5.0%, p=0.033), headache (50% vs. 39%, p=0.034), and abdominal pain (14% vs. 8.3%, p=0.043) were more frequent in patients from the pre-Omicron phase compared to the Omicron phase. In contrast, muscle ache was more common in the Omicron phase than pre-Omicron phase (35% vs 25%, p=0.029).

**Table 5.** Prevalence of symptoms among PCR-confirmed SARS-CoV-2 infected adult patients during Omicron and pre-Omicron phases.

| Characteristic | Proportions |  |  |  | N | Univariate |  |  | Multivariable |  |  |
| --- | --- | --- | --- | --- | --- | --- | --- | --- | --- | --- | --- |
|  | Overall, N = 447<sup>1</sup> | Omicron phase, N = 302<sup>1</sup> | Pre-omicron phase, N = 145<sup>1</sup> | p-value<sup>2</sup> |  | OR<sup>3</sup> | 95% CI<sup>3</sup> | p-value | OR<sup>3</sup> | 95% CI<sup>3</sup> | p-value |
| **Fever** | 77 (17%) | 58 (19%) | 19 (13%) | 0.11 | 447 |  |  |  |  |  |  |
| No |  |  |  |  |  | Ref. | — |  | Ref. | — |  |
| Yes |  |  |  |  |  | 1.58 | 0.91, 2.82 | 0.11 | 2.33 | 1.23, 4.66 | **0.012** |
| **Cough** | 264 (59%) | 162 (54%) | 102 (70%) | **<0.001** | 447 |  |  |  |  |  |  |
| No |  |  |  |  |  | Ref. | — |  | Ref. | — |  |
| Yes |  |  |  |  |  | 0.49 | 0.32, 0.74 | **<0.001** | 0.36 | 0.21, 0.60 | **<0.001** |
| **Fatigue** | 32 (7.2%) | 11 (3.6%) | 21 (14%) | **<0.001** | 447 |  |  |  |  |  |  |
| No |  |  |  |  |  | Ref. | — |  | Ref. | — |  |
| Yes |  |  |  |  |  | 0.22 | 0.10, 0.47 | **<0.001** | 0.18 | 0.07, 0.41 | **<0.001** |
| **Loss of smell** | 16 (3.6%) | 8 (2.6%) | 8 (5.5%) | 0.13 | 447 |  |  |  |  |  |  |
| No |  |  |  |  |  | Ref. | — |  | Ref. | — |  |
| Yes |  |  |  |  |  | 0.47 | 0.17, 1.29 | 0.13 | 0.45 | 0.10, 1.97 | 0.3 |
| **Loss of taste** | 30 (6.7%) | 15 (5.0%) | 15 (10%) | **0.033** | 447 |  |  |  |  |  |  |
| No |  |  |  |  |  | Ref. | — |  | Ref. | — |  |
| Yes |  |  |  |  |  | 0.45 | 0.21, 0.96 | **0.037** | 0.81 | 0.27, 2.51 | 0.7 |
| **Sore throat** | 64 (14%) | 47 (16%) | 17 (12%) | 0.3 | 447 |  |  |  |  |  |  |
| No |  |  |  |  |  | Ref. | — |  | Ref. | — |  |
| Yes |  |  |  |  |  | 1.39 | 0.78, 2.58 | 0.3 | 1.44 | 0.75, 2.86 | 0.3 |
| **Headache** | 190 (43%) | 118 (39%) | 72 (50%) | **0.034** | 447 |  |  |  |  |  |  |
| No |  |  |  |  |  | Ref. | — |  | Ref. | — |  |
| Yes |  |  |  |  |  | 0.65 | 0.44, 0.97 | **0.035** | 0.47 | 0.29, 0.74 | **0.001** |
| **Muscle ache** | 142 (32%) | 106 (35%) | 36 (25%) | **0.029** | 447 |  |  |  |  |  |  |
| No |  |  |  |  |  | Ref. | — |  | Ref. | — |  |
| Yes |  |  |  |  |  | 1.64 | 1.06, 2.58 | **0.030** | 1.59 | 0.94, 2.76 | 0.089 |
| **Joint pain** | 42 (9.4%) | 26 (8.6%) | 16 (11%) | 0.4 | 447 |  |  |  |  |  |  |
| No |  |  |  |  |  | Ref. | — |  | Ref. | — |  |
| Yes |  |  |  |  |  | 0.76 | 0.40, 1.49 | 0.4 | 0.69 | 0.30, 1.61 | 0.4 |
| **Abdominal pain** | 46 (10%) | 25 (8.3%) | 21 (14%) | **0.043** | 447 |  |  |  |  |  |  |
| No |  |  |  |  |  | Ref. | — |  | Ref. | — |  |
| Yes |  |  |  |  |  | 0.53 | 0.29, 1.00 | **0.046** | 0.37 | 0.18, 0.76 | **0.007** |
| **Diarrhoea** | 24 (5.4%) | 14 (4.6%) | 10 (6.9%) | 0.3 | 447 |  |  |  |  |  |  |
| No |  |  |  |  |  | Ref. | — |  | Ref. | — |  |
| Yes |  |  |  |  |  | 0.66 | 0.29, 1.56 | 0.3 | 0.69 | 0.27, 1.82 | 0.4 |
| **Shortness of breath** | 28 (6.3%) | 20 (6.6%) | 8 (5.5%) | 0.7 | 447 |  |  |  |  |  |  |
| No |  |  |  |  |  | Ref. | — |  | Ref. | — |  |
| Yes |  |  |  |  |  | 1.21 | 0.54, 2.99 | 0.7 | 2.65 | 0.88, 8.76 | 0.094 |
| **Chest pain** | 73 (16%) | 46 (15%) | 27 (19%) | 0.4 | 447 |  |  |  |  |  |  |
| No |  |  |  |  |  | Ref. | — |  | Ref. | — |  |
| Yes |  |  |  |  |  | 0.79 | 0.47, 1.34 | 0.4 | 0.83 | 0.46, 1.53 | 0.5 |
| **Runny nose** | 104 (23%) | 65 (22%) | 39 (27%) | 0.2 | 447 |  |  |  |  |  |  |
| No |  |  |  |  |  | Ref. | — |  | Ref. | — |  |
| Yes |  |  |  |  |  | 0.75 | 0.47, 1.18 | 0.2 | 1.10 | 0.65, 1.90 | 0.7 |
| **Pneumonia** | 24 (5.4%) | 15 (5.0%) | 9 (6.2%) | 0.6 | 447 |  |  |  |  |  |  |
| No |  |  |  |  |  | Ref. | — |  | Ref. | — |  |
| Yes |  |  |  |  |  | 0.79 | 0.34, 1.92 | 0.6 | 0.35 | 0.11, 1.10 | 0.071 |
| **Sex** |  |  |  | 0.2 | 447 |  |  |  |  |  |  |
| Female | 232 (52%) | 151 (50%) | 81 (56%) |  |  | Ref. | — |  | Ref. | — |  |
| Male | 215 (48%) | 151 (50%) | 64 (44%) |  |  | 1.27 | 0.85, 1.89 | 0.2 | 1.38 | 0.89, 2.16 | 0.15 |
| **Age** | 31 (24, 39) | 31 (25, 38) | 32 (23, 39) | 0.8 | 447 | 1.00 | 0.98, 1.02 | 0.7 | 0.99 | 0.97, 1.02 | 0.5 |
| **COVID-19 vaccinated** |  |  |  | 0.5 | 447 |  |  |  |  |  |  |
| No | 310 (69%) | 206 (68%) | 104 (72%) |  |  | Ref. | — |  | Ref. | — |  |
| Yes | 137 (31%) | 96 (32%) | 41 (28%) |  |  | 1.18 | 0.77, 1.84 | 0.5 | 1.35 | 0.83, 2.22 | 0.2 |

<sup>1</sup> n (%); Median (IQR)
<sup>2</sup> Pearson's Chi-squared test; Wilcoxon rank sum test
<sup>3</sup> OR = Odds Ratio, CI = Confidence Interval

Controlling for age, sex and COVID-19 vaccination status in a multivariable analysis, cough (OR 0.37 [CI 0.22 – 0.61], p<0.001), fatigue (OR 0.20 [CI 0.08 – 0.48], p<0.001), headache (OR 0.47 [CI 0.29 – 0.79], p=0.001), and abdominal pain (0R 0.38 [CI 0.18 – 0.78) were independently associated with pre-Omicron than Omicron SARS-CoV-2 PCR positive diagnosis (**figure 2 and table 5)**. Conversely, fever was independently associated with Omicron than pre-Omicron SARS-CoV-2 PCR positive diagnosis (OR 2.46 [CI 1.29 – 4.97]) (**figure 2 and table 5)**.

**Figure 2.**
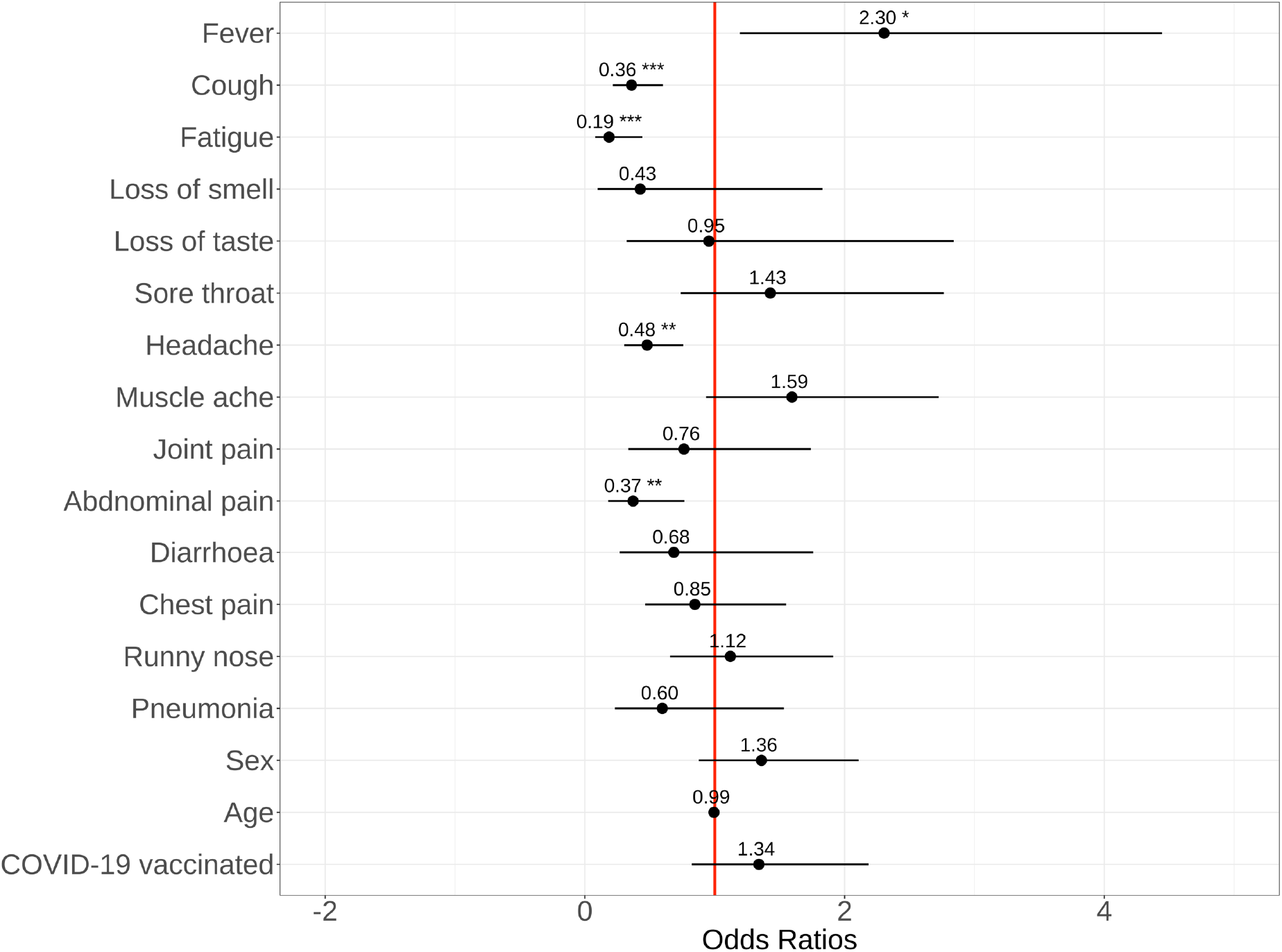
Clinical symptoms associated with PCR-confirmed SARS-CoV-2 infection. Differences in clinical symptoms of PCR-confirmed SARS-CoV-2 infected adults during the Omicron and Pre-Omicron phases (n=447).

## DISCUSSION

In a setting where there is a high burden of presentations with infectious disease, we found that the symptoms associated with SARS-CoV-2 Omicron infection have become considerably less distinct, differing significantly from those infections with pre-Omicron variants (predominantly Delta). Indeed fever, which is common to many infectious presentations ^11,12^, was most prevalent among presumed Omicron infected patients, while headache, cough, fatigue, and abdominal pain were significantly more prevalent among pre-Omicron cases.

Our data showing a different symptom profile associated with Omicron infection is consistent with studies elsewhere ^21^ with headache being prominent in three other studies from the UK ^21-23^. However, we observed high odds for presenting with fever in presumed Omicron-infected patients than pre-Omicron patients, in contrast with the two studies in the UK ^21,23^. The main differences between the Malawi study and the UK studies are age and prevalence of Omicron sub-lineages, with Malawi cohort being a younger population and having predominantly BA.1 at time of sampling. BA.1 is associated with a different symptom profile than BA.2 ^22^. Together, our findings and those of others suggest that the clinical case definition of COVID-19 used for testing and surveillance may need to be revised to maintain case ascertainment.

Data from Malawi and elsewhere has shown that the Omicron variant has presented with less severe disease, hospitalisation and deaths, than the pre-Omicron VOCs ^1,24^. The Omicron variant have been shown to be less capable of transition from the upper to lower respiratory tract infection ^25^, and this could potentially contribute to the low incidence of severe disease. However, data in non-immunised populations in Hong Kong indicate that Omicron is not intrinsically mild ^26,27^, suggesting that immune response or past exposure could be an important determinant of this low severity. Data from South Africa has shown that high SARS-CoV-2 seroprevalence has been associated with low number of deaths and hospitalisation attributed to the Omicron variant ^24^. In Malawi, seroprevalence data has shown that more than 70% of the population had anti-SARS-CoV-2 receptor binding domain (RBD) antibodies before the Omicron variant pandemic wave ^5^, reaching over 90% at the peak of the Omicron wave in January 2022 (unpublished). In line with previous findings ^28^, COVID-19 vaccination was not associated with a reduced risk of PCR-confirmed SARS-CoV-2 infection, especially during the Omicron wave. It is therefore plausible that the altered clinical presentation observed in our study could also be attributed to pre-existing immunity from prior SARS-CoV-2 exposure.

Furthermore, our findings align with the temporal dynamics of the COVID-19 pandemic waves in Malawi and the region ^1,5,24^. A high SARS-CoV-2 prevalence of 30-50% among patients presenting to primary healthcare at the peak of the three pandemic waves, is consistent with high reported national COVID-19 cases during the same period ^1^. Furthermore, consistent with genomic surveillance ^15,29^, our sentinel surveillance correctly identified the VOCs driving the local pandemic waves. Due to the consistency in our sampling over time, we were able to provide real-time data to aid public health response in Malawi, especially on the identification of VOCs driving community transmission (unpublished). Collectively, this indicates that sentinel surveillance backed up by diagnostics and genomics data could be an early warning system for national pandemic response in resource-limited settings, considering that by the time hospitalisations are rising it is already too late to intervene effectively.

Our study had several limitations. Firstly, the study was conducted in urban Blantyre and findings may not be generalisable to rural settings. Secondly, our study cohort (median age 28 years [IQR 21-38]) was not fully representative of the population structure in Malawi (median age 17.5 years) ^30^. Thirdly, since our genomic surveillance was limited to a subset of samples with low PCR CT values, this approach biases our identification of variants to those causing high viral burden infections at time of recruitment.

In conclusion, our study demonstrates changes in clinical symptoms overtime, aligned to infecting variant, indicating that case definitions of COVID-19 need constant monitoring and revision to match SARS-CoV-2 evolution to maintain its relevance for institutional and national testing policies. This study also highlights the importance and utility of sentinel surveillance in low-resourced settings to aid timely public health response against the COVID-19 pandemic and future pandemics.

## Data Availability

All data produced in the present study are available upon reasonable request to the authors

## NOTES

### Author contributions

Conceptualization of study: RSH, NF, TS, MGC, KCJ

Study design: RSH, NF, TS, MGC, KCJ

Data collection: SN, CT, FK, AA, NM

Laboratory testing: CA, KB, JC

Data analysis: PMA, KCJ

Accessed and verified the underlying data: HWT, KCJ

Study management: HWT, MG, HT, VN, DM, EA

Manuscript writing: MGC, HWT, KCJ, NF

Manuscript review and approval: All authors

## ACKNOWLEDGEMENTS

The authors thank all participants whose samples are used in this study. We thank the MLW Clinical Research Support Unit (CRSU) team for supporting this study.

## DISCLAIMER

The findings and conclusions in this report are those of the authors and do not necessarily represent the official position of the funders.

## Supplementary Material

**Supplementary Table 1.**

| Characteristic | Overall, N = 35<sup>1</sup> | Omicron phase, N = 26<sup>1</sup> | Pre-omicron phase, N = 9<sup>1</sup> | p-value<sup>2</sup> |
| --- | --- | --- | --- | --- |
| **Sex** |  |  |  | 0.5 |
| Female | 16 (46%) | 13 (50%) | 3 (33%) |  |
| Male | 19 (54%) | 13 (50%) | 6 (67%) |  |
| **Age group** |  |  |  | 0.3 |
| <13 years | 28 (80%) | 22 (85%) | 6 (67%) |  |
| 13-17 years | 7 (20%) | 4 (15%) | 3 (33%) |  |
| **Asthma** | 1 (2.9%) | 1 (3.8%) | 0 (0%) | >0.9 |
| <sup>1</sup> n (%) |  |  |  |  |
| <sup>2</sup> Fisher's exact test |  |  |  |  |

**Supplementary Figure 1.**
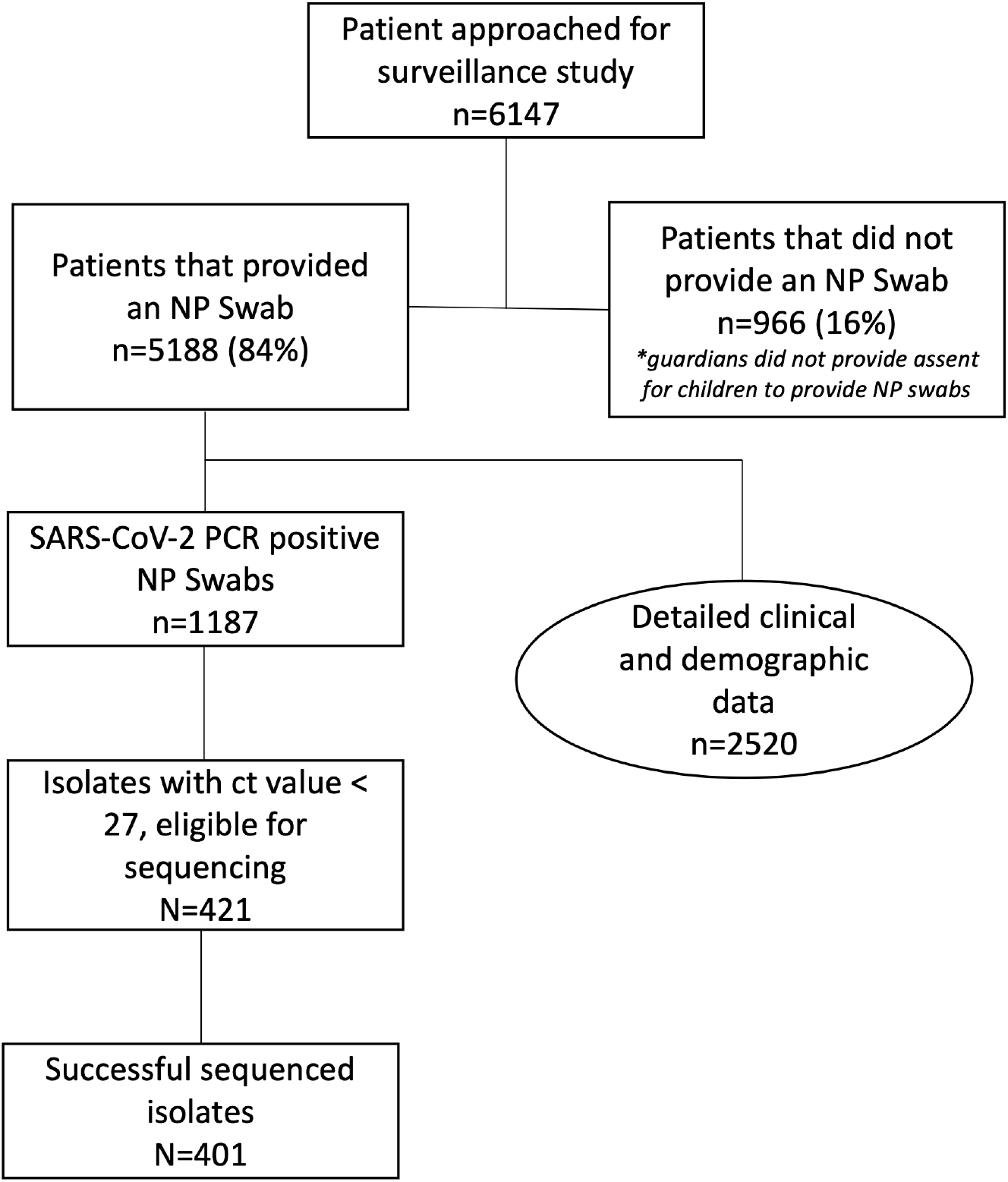
Flowchart of participant enrolment and testing in two primary healthcare facilities in Blantyre City, Malawi, November 2020 – March 2022.

